# Brain Dynamics of Persistent Postural-Perceptual Dizziness at Rest Reveal Alterations in Multisensory and Motor Areas

**DOI:** 10.64898/2026.10.26.26366555

**Authors:** Cristina Concetti, Anna Wyss, Salome Häuselmann, Irene Lozzi, Tatiana Brémovà-Ertl, Hassen Kerkeni, Georgios Mantokoudis, Nicolas Gninenko, Michael Mouthon, Maria Giulia Preti, Dimitri Van De Ville, Selma Aybek

## Abstract

**Background:** Persistent Postural-Perceptual Dizziness (PPPD) is a chronic functional disorder with unsteadiness and postural instability, attributed to altered multisensory integration and visual dependence. fMRI studies report altered connectivity in several regions, however, dynamic connectivity remains unexplored.

**Objectives:** We characterized resting-state brain dynamics in 17 patients with PPPD and 20 matched healthy controls (HC), and their relationship to clinical and postural measures.

**Methods:** Co-Activation Patterns (CAPs) were derived from resting-state fMRI scans for four seeds relevant to multisensory integration, motor output and vestibular processing – right temporo-parietal junction (rTPJ), caudate nucleus, hippocampus and operculo-parietal area 2 (OP2) – and their temporal metrics compared between groups. Partial Least Squares Correlation (PLSC) related CAPs to mood, vertigo, stress, and visual dependence for gait and postural control.

**Results:** Patients with PPPD showed altered dynamics seeded from rTPJ (decrease of a salience-coupled CAP, increased entries into a visual/dorsal-attention-coupled CAP), caudate nucleus (increased time in a default mode-coupled CAP, decrease of a salience/visual-coupled CAP) and hippocampus (increase of a default-mode-coupled CAP), but not OP2. PLSC yielded two significant latent components: the first linked rTPJ, OP2 and hippocampal CAPs sharing visual network co-deactivation to stress, mood, vertigo and visual dependence for gait; the second linked all caudate and two rTPJ CAPs to postural sway with eyes closed (visual dependence). Individual brain-behavior scores correlated in both groups.

**Conclusion:** Resting-state dynamics are altered in multisensory and motor regions in PPPD, but preserved in OP2, the core vestibular cortex. Brain-behavior organization appears shared across groups, with shifted dynamics in patients.

**Graphical abstract:** Curved arrows indicate dynamic functional connectivity patterns (Co-Activation Patterns) differing between patients with PPPD and HC (the direction is indicated by the small straight arrows). Dashed arrows indicate latent components of brain-behavior coupling identified through PLSC analysis. (rTPJ: right temporo-parietal junction; OP2: parietal operculum area 2; AP: antero-posterior).

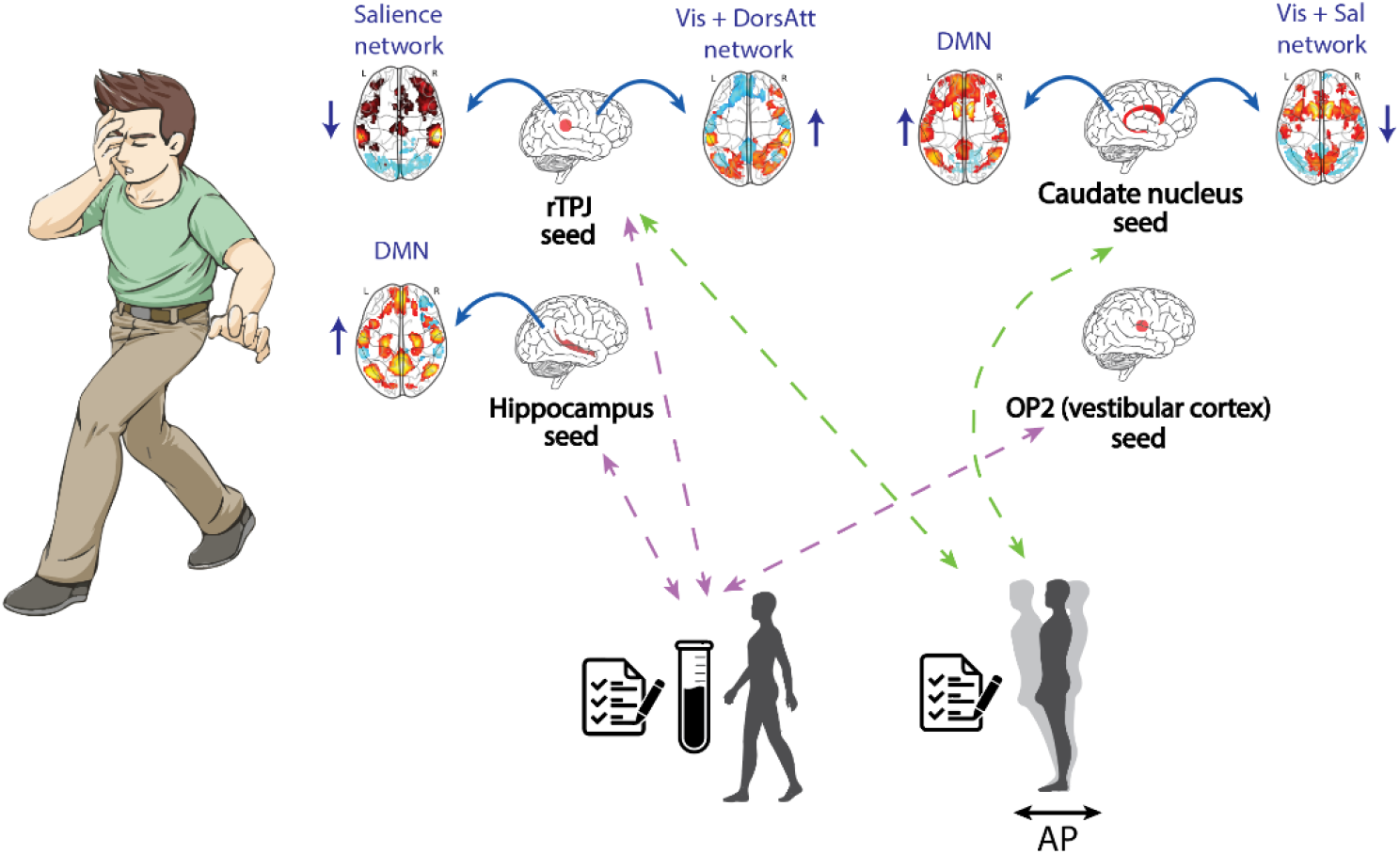

## 1. Introduction

Persistent Postural-Perceptual Dizziness (PPPD) is a functional vestibular disorder that often takes a chronic course[1,2]. Patients experience feelings of swaying and unsteadiness on most days, which are often exacerbated by upright posture, complex visual stimuli, and active or passive motion[3]. Due to its recent definition (in 2017, by the International Classification of Vestibular Disorders)[3], incidence estimates are still uncertain, but it is recognized as one of the most common causes of chronic dizziness[4]. The therapeutic approach to this disorder is Vestibular Physical Therapy, aiming at postural retraining[5]. PPPD is closely related to the diagnostic category of Functional Neurological Disorders (FND), which are characterized by variable neurological symptoms (motor, sensory, cognitive) or functional seizures (i.e., seizure-like attacks without epileptiform activity)[6]. Research has shown that FND and PPPD display alterations at the behavioral and motor level, and in brain activation and functional connectivity[7–9]. They also present psychiatric comorbidities, most often symptoms of anxiety and depression[1,2,10].

PPPD is thought to develop after the recovery from an initial vestibular insult, or an insult that causes acute balance disruption[11]. This first episode of dizziness might induce adaptive adjustments, such as the down-weighing of vestibular inputs and the up-weighing of visual inputs for postural control. However, a failure to recalibrate these inputs after the resolution of the initial insult can lead to long-term maladaptive over-reliance on visual stimuli and an excessive explicit attentional control on posture, as observed in patients with PPPD[1,11].

Structural neuroimaging studies of PPPD have generally reported a reduction of grey matter in multisensory vestibular processing regions, and an increase in regions regulating mood[8], which has been suggested to reflect the implication of affective symptoms in the maintenance of this disorder. Functional neuroimaging studies with resting-state fMRI have reported alterations in the connectivity between visual and vestibular networks, multisensory regions, such as the precuneus, as well as frontal regulatory regions, and in hippocampal and cerebellar circuits[8,11–13].

Despite recent advancements aimed at identifying alterations in functional connectivity, behavioral and clinical aspects, and their role in the disorder, the involvement of different factors in its pathophysiology is still unclear. To fill this gap, we have conducted a multilevel study investigating self-reported, clinical, neuropsychological, biological, and behavioral variables relevant to PPPD, in the same cohort of patients and matched healthy control subjects (HC). We have previously reported clinical data on group differences in these variables[14], and we here set out to add advanced analysis of resting state fMRI, allowing us to test for dynamic changes. Clinical data revealed [14] an increase in affective symptoms and perceived stress in patients with PPPD, but no differences in biological stress markers or cognition compared to HC. Additionally, we reported that patients with PPPD present a strong dependence on vision for gait and postural control – with slowing locomotion and increased instability in the anteroposterior direction upon closing their eyes – while challenges to other sensory systems did not elicit the same effect. This allowed us to shed more light on the contribution of these factors in our sample but raises the question on their relationship with brain activity signatures.

Therefore, here we investigate the brain activity of patients with PPPD and HC at rest through fMRI, and its relationship with self-reported and behavioral variables. We employ a dynamic functional connectivity analysis: seed-based Co-Activation Patterns (CAPs)[15,16]. Classical seed-based functional connectivity examines correlations between the seed time course and the rest of the brain across the entire recording time, thus treating these as fixed or static. However, it has been shown that functional connectivity can vary dynamically over time while recording the brain at rest[17], and CAPs analysis has been developed to capture these transient states. The CAPs approach has been employed so far in several clinical populations (e.g., [18–21]), but not in PPPD. This disorder can involve spontaneous and transient symptom fluctuations, lasting for seconds[3], thus an analysis approach with the same time scale might uncover relevant temporary fluctuations in brain activity. Additionally, dynamic pattern fluctuations can reveal differences in network reconfigurations that static connectivity would miss by averaging.

We have chosen to focus our CAPs analysis on four seed regions that have relevance for this disorder: the right temporo-parietal junction (rTPJ), the caudate nucleus, the hippocampus, and the parietal operculum area 2 (OP2). This multi-seed approach is based on the fact that neuroimaging research on PPPD so far highlights the involvement of multiple regions and networks[12,13]. The rTPJ is involved in multisensory integration, including that of visual and vestibular stimuli[22], and in the sense of body ownership and self-localization[23,24]. It has been repeatedly implicated in FNDs[7] and associated to alterations in the sense of agency[25] (the feeling of being in control of one’s actions), which is also altered in these disorders (e.g., [26]). However, its role in the functional connectivity in PPPD is not known. The caudate nucleus is an important region for motor planning and sensorimotor integration[27,28], and its functional connectivity with the insula (part of the salience network) has been found to be altered in PPPD[29], however its wider involvement remains to be elucidated. The hippocampus is known for its central role in memory processes and spatial navigation. Previous studies have reported a reduced grey matter volume of this structure[30], and altered connectivity of the hippocampus with several brain regions[31] in patients with PPPD compared to HC. Finally, the OP2 has been identified as the core human vestibular cortex[32,33] and previous studies have reported mixed results on its connectivity in PPPD[31,34], calling for additional investigations of this area. We hypothesized that the rTPJ, caudate nucleus and hippocampus will show alterations in dynamic functional connectivity.

We have next explored the relationship between brain dynamics derived from these seed regions, i.e., CAPs, and clinical and behavioral variables, through Partial Least Squared Correlation (PLSC)[35], a multivariate correlation analysis, which can provide insights on the functional role of identified brain signatures, namely which alterations in dynamic functional connectivity are related to postural instability.

## 2. Methods

### 2.1 Participants

40 participants were enrolled in this study between 2023 and 2024. 20 patients with PPPD were recruited at the Dizziness Center and the Psychosomatic Unit of the Bern University Hospital and 20 age- and sex-matched HC were recruited through advertisement and word-of-mouth. Patients with PPPD were diagnosed by a certified neurologist in accordance with the ICD-11 (International Classification of Diseases, 11th revision)/Bárány Society diagnostic criteria[3]. All participants were selected to be above 16 years of age, capable of judgement and willing to participate in the study. Exclusion criteria were: severe comorbid psychiatric disorders (e.g. psychosis or suicidality), neurological conditions (e.g., epilepsy), history of drug or alcohol abuse, past brain surgery; inability to follow the study protocol, contraindications for MRI, e.g., implanted metal clips or medical devices. Additionally, for female participants, pregnancy, breastfeeding, or the intention to become pregnant also constituted exclusion criteria.

The study was approved by the Ethical Committee of the Canton of Bern (SNCTP000004529,2020-02283) and registered on ClinicalTrials.gov (NCT05086380). The study was conducted according to the Declaration of Helsinki and Good Clinical Practice, at the University Hospital of Bern, with neuroimaging being carried out at the Translational Imaging Center SITEM-Insel. All participants provided written informed consent and were reimbursed for travel expenses.

Two participants were excluded from MRI data analysis due to corrupted data, and one was excluded due to excessive motion (FD>0.5 mm, also see Section 2.5), resulting in a final sample of 20 HC and 17 patients with PPPD.

### 2.2 Demographic and clinical data

Demographic information, such as age, sex, smoking and medication use was collected. Participants also underwent a clinical and self-report evaluation. Depressive symptomatology was quantified using the BDI-I (Beck’s Depression Inventory)[36], while anxiety was assessed through STAI (State and Trait Anxiety Inventory)[37]. To capture both depressive and anxious symptomatology – often present in patients with PPPD[2] – we calculated the mood score as the average of the individual z-scored depression and trait anxiety scores, similarly to our previous work[38]. Perceived stress was measured via the PSS (Perceived Stress Scale)[39]. To evaluate vestibular-related symptomatology and its impact on daily activities, participants completed the NVI (Neuropsychological Vertigo Inventory)[40]. Clinical and behavioral results used here to investigate brain-behavior relationships through PLSC analysis are described and discussed in more detail in[14].

### 2.3 Salivary Stress Markers

Salivary cortisol and α-amylase were sampled from participants using cotton swabs (Salivette CollectionDevices, Sarstedt) held in the mouth for ∼1 min per sample. All saliva samples were centrifuged at room temperature for 10 min at 3500 rpm and subsequently stored at −20°C. Salivary cortisol and α-amylase concentrations were determined using commercial enzyme immunoassays (Salimetrics, High Sensitivity Salivary Cortisol Enzyme Immunoassay kit, 1-3002; Salimetrics, Salivary Alpha-Amylase Kinetic Enzyme Assay Kit, 1-1902), following the manufacturer’s protocols. Further details on the methods and data analysis related to salivary stress markers are described and discussed in the companion article[14].

### 2.4 Gait and Postural Assessments

Gait parameters were acquired using the GAITRite walkway system (CIR Systems Inc., Franklin, NJ, USA), an electronic carpet able to record gait characteristics. Briefly, participants walked barefoot under several conditions, including normal walking (i.e., self-selected speed), and walking with eyes closed. Gait velocity was extracted and averaged across valid trials. Gait velocity Δ was calculated as the difference in gait velocity with eyes open and eyes closed.

Static postural control was evaluated using posturography (Kistler force plate, NeuroPlatform software). Participants underwent a standardized protocol, including eyes open and eyes closed conditions. Safety measures (light touch, hand support, or examiner assistance) were applied if necessary to prevent falls. Center of pressure (CoP) displacement signals were recorded and CoP swaypath in the mediolateral (X) and anteroposterior (Y) directions were extracted. Postural sway Δ in the two directions was calculated as the difference in postural sway with eyes open and eyes closed.

More information on the acquisition and data analysis of gait and postural assessments can be found in[14].

### 2.5 MRI data acquisition and preprocessing

Structural and functional MRI volumes were acquired using a 3 Tesla scanner (Magnetom Prisma, Siemens, Germany). To minimize head motion, participants’ heads were stabilized using foam pads. They were instructed to remain still, stay awake, and to fixate a white cross presented on a black background. The structural scan consisted of a sagittal-oriented T1-weighted 3D-magnetization-prepared rapid acquisition gradient echo (MPRAGE) sequence with 1 mm isotropic voxel size (repetition time TR=2330 ms, echo time TE=3.03 ms, inversion time TI=1100 ms, flip angle FA= 8°, matrix = 256 × 256 × 176, acquisition time = 5:27 min). The resting-state functional scan consisted of a whole-brain, inter-leaved, multi-slice BOLD echo-planar imaging (EPI) sequence with an isotropic voxel size of 2.2 mm (TR=1300 ms, TE=37 ms, FA=52°, matrix size = 104 × 104 × 60, acquisition time = 6:30 min), yielding a total of 300 functional volumes. Participants were instructed to keep their eyes open and look at a white fixation cross.

MRI data were preprocessed using SPM12 (Wellcome Centre for Human Neuroimaging,UCL, London) in MATLAB (R2022b, MathWorks Inc., Natick, USA). Functional volumes were initially realigned, unwarped (two participants were excluded due to corrupted files), and co-registered to the corresponding structural T1-weighted image. The structural images were segmented into white matter, grey matter and CSF (cerebrospinal fluid) and an individual brain mask was created. Single-voxel time series of functional images were detrended (average signal) and corrected for nuisance contributions by regressing out the average white matter and CSF signals, the six motion parameters (three translational and three rotational), the global signal, and low-frequency trends (constant, linear and quadratic). A high-pass filter with a cutoff frequency of 0.01 Hz was then applied to remove low-frequency drifts. Lastly, functional volumes were warped into MNI standard space and spatially smoothed using a Gaussian kernel with a full width at half maximum (FWHM) of 5 mm. Functional images were assessed for excessive head motion using the framewise displacement (FD) threshold of 0.5 mm.

### 2.6 Co-Activation Patterns analysis

To identify transient patterns of connectivity, CAPs analysis starts with selecting volumes (i.e., time points) at which the seed region is highly active. This has been shown to yield a map that recapitulates the functional connectivity of the seed. Then, by clustering highly co-active and co-deactive voxels, distinct spatial patterns can be identified[41].

Co-Activation Patterns analysis was performed using the TB-CAPs Toolbox[15] (https://github.com/MIPLabCH/TbCAPs). Masks for seed regions were bilateral for all except for the right temporo-parietal junction (rTPJ). Masks for the caudate nucleus and hippocampus were generated in SPM12 using the wfupickatlas toolbox (https://www.nitrc.org/projects/wfu_pickatlas) with the AAL atlas, while the mask for the OP2 region was obtained using the Julich Brain Atlas with siibra explorer (https://atlases.ebrains.eu/viewer). The mask for the rTPJ was obtained using the wfupickatlas toolbox in SPM12, as a sphere of 10 mm centered at [62 –34 30] in the MNI (Montreal Neurological Institute) space, as in our previous work[42].

For CAPs analysis, the preprocessed fMRI data was used. The time series of the seed region was z-scored and thresholded at 0.84 SD to identify frames of high activity in the BOLD signal of the seed. Frames exceeding an FD threshold of 0.5 mm, together with the adjacent (one previous and one subsequent) frames, were scrubbed from the data. At this stage, we also verified that the number of retained (highly active) frames was comparable for the two groups (**Suppl. Fig.1a-d**, left, and **Suppl. Table 1**). An additional principal component analysis (PCA) step was applied at this stage to reduce the high dimensionality of the data and its result used as input for the next step, as in our previous work[43]. To determine the optimal number of clusters K that define the Co-Activation Patterns, we performed consensus clustering on the PCA scores of HCs, used as a reference population. The optimal number of clusters was identified based on consensus clustering metrics: the stability measure (1-PAC, where PAC is the Percentage of Ambiguously Clustered pairs), and the visual inspection of consensus matrices (**Suppl. Fig1a-d**, middle and right panels) [44]. The optimal number of clusters was also chosen as K>2 to avoid a trivial solution constituted by only a pattern and its opposite, as recommended with this method[41]. The fMRI volumes were then clustered into the identified optimal number K of distinct states (CAPs), using the k-means algorithm (50 repetitions). The individual CAPs were then reconstructed by reversing the dimensionality reduction and spatially z-scored and thresholded (1.7 SD), to show both positive and negative contributions. These voxels within each CAP were mapped onto functional networks by computing their spatial overlap with networks from the Yeo 17-networks atlas[45], after removing the seed region (a complete list of regions contributing to each CAP can be found in **Suppl. Table 2**). Next, the retained frames from the patients with PPPD’ population were assigned to the most similar CAP based on spatial correlation with a threshold above the 5th percentile of HC frames[15]. This generally resulted in <10% of unassigned frames in the PPPD group (**Suppl. Table 1**). CAPs brain visualizations were generated using the AtlasReaderpackage (https://github.com/miykael/atlasreader) in Python (version 3.12.7).

To analyze the temporal characteristics of the identified CAPs, we computed three temporal metrics as in previous work[21,43]: relative occurrence, relative entries and average duration. The relative occurrence is the overall number of volumes assigned to a given pattern, normalized by the number of retained frames (which can slightly differ across individuals), indicating the total time spent by the brain in a given state (also called dwell time). Relative entries are calculated as the number of times a CAP is entered, normalized by the number of retained frames, indicating how often the brain enters a specific pattern, i.e. CAP frequency. The average duration, calculated as the mean number of consecutive frames assigned to the same pattern and multiplied by the TR, reflects how long a brain state remains stable once it is entered.

### 2.7 PLSC analysis

Partial Least Squares Correlation (PLSC) is a method commonly used in neuroimaging to analyze associations between two datasets. It searches for Latent Components (LCs, linear combinations of the original variables) that express the largest amount of information common to the two sets of data (maximal covariance), in this case brain data on one side, and clinical and behavioral data on the other. Its outputs include the *salience* of each variable, indicating the weight that variable holds in constructing the LC, and the *loading* of each variable, indicating its correlation once the variable is projected onto the component.

Grouped PLSC analysis was implemented using the publicly available MATLAB-based PLS toolbox (https://github.com/FND-ResearchGroup/myPLS_SL), similarly to our previous work[21,43]. Due to some missingness of clinical and behavioral data, we adopted a strictly conservative approach for this analysis and only included participants with complete datasets, resulting in a final sample of 17 HC and 16 patients with PPPD. Among the several available clinical and behavioral variables, we selected those which were *a priori* considered most meaningful, trying to capture a comprehensive multi-level characterization relevant to PPPD: the area under the curve of the awakening cortisol response (‘Cortisol AUC’)[46], the concentration of salivary α-amylase (U/mL, ‘Amylase’), a mood score calculated as the averaged combined z-score of the BDI and STAI-T, the total score of the NVI, the PSS, the difference of gait velocity with eyes open and closed (‘Gait velocity Δ’), the difference of postural sway in the medio-lateral and antero-posterior directions with eyes open and closed (‘Postural Sway ML Delta’ and ‘Postural Sway AP Delta’, respectively). The selection of these variables was not based on previous knowledge on group differences, to avoid circularity. These variables were entered into the ‘behavioral’ or ‘Y’ matrix of the PLSC analysis, while the relative occurrence metric for all CAPs constituted the ‘brain’ or ‘X’ matrix. Relative occurrence was selected among the available temporal metrics, as it is by definition a combination of entries and duration, thus capturing most of the temporal features of CAPs, while reducing the risk of collinearity of the input ‘brain’ data. All CAPs were included, independently of the previous findings on group differences, to avoid circularity in the analysis. A selection of ‘brain’ and ‘behavioral’ variables was also necessary to avoid the risk of overfitting that arises when the dimensionality of variables is greater than the sample size[47].

### 2.8 Statistical analysis

Statistical comparisons of demographic and clinical data were conducted with parametric and non-parametric t-tests based on normality and variance assumptions as appropriate, using JASP (Version 0.95.4). The temporal characteristics of CAPs were analyzed using simple linear models using Rstudio (Posit, version 2025.09.1), for each metric and across all CAPs (temporal metric ∼ group*CAP), as preliminary model fitting showed that random effects were close to zero. For post-hoc comparisons, a global FDR correction (Benjamini-Hochberg procedure) was applied to account for testing across the different CAPs and the 3 different metrics. For PLSC analysis, the statistical significance of the identified LC was tested through permutation (5000 permutations), and the robustness of the saliences and loadings through bootstrapping (5000 bootstrap samples, i.e. resamples with replacement). Details of statistical analyses are presented in figure captions.

## 3. Results

### 3.1 Participants’ demographics and clinical characteristics

The PPPD group and HC group were matched in age and sex, but more patients with PPPD presented intake of psychotropic medication compared to HC (**Table 1**). Patients with PPPD also showed significantly higher scores for self-reported symptomatology of depression and a trend for increased trait anxiety, with no significant differences in state anxiety. The perceived stress was significantly higher in patients with PPPD, while the impact of vertigo (NVI) showed only a trend (**Table 1**). Further characterization of this cohort is described and discussed in depth in the companion article[14]. In brief, overall, patients with PPPD showed increased symptoms of depression and stress compared to HC.

**Table 1.** Main demographic and clinical characteristics of the study population. . The PPPD and HC groups were well matched for age and sex. They differ in their scores for depression, perceived stress, and mood (composite score), but not in their anxiety scores (with a trend for increased trait anxiety) and present a trend for increased impact of vertigo (NVI). The combined Mood Score was calculated as the average of the z-scored individual values of depression and state anxiety to obtain a composite score. (§ = Mann-Whitney U-test; ° = Chi-squared test; no symbol = Student’s t-test; * = significant with 0.05>p≥0.01 ** = significant with p<0.01; M = male, F = female).

|  | PPPD (M±SD) | HC (M±SD) | p |
| --- | --- | --- | --- |
| ***N*** | 18 | 20 |  |
| ***Age*** | 44.8 ± 15.6 | 41.6 ± 15.8 | p=0.59§ |
| ***Sex*** | 9 M – 9 F | 7 M – 13 F | p=0.35° |
| ***Intake of psychotropic medication*** | 7 | 1 | p=0.01°* |
| ***Smokers*** | 1 | 1 | p=0.94° |
| ***Depression (BDI)*** | 9.83 ± 7.79 | 4.00 ± 4.03 | p<0.01§** |
| ***Anxiety – trait (STAI-T)*** | 40.55 ± 11.86 | 34.25 ± 8.92 | p=0.07 |
| ***Anxiety – state (STAI-S)*** | 38.11 ± 10.83 | 32.05 ± 6.77 | p=0.10 |
| ***Mood score*** | 0.41 ± 1.02 | -0.36 ± 0.62 | p<0.01** |
| ***Vertigo (NVI)*** | 39.78 ± 15.89 | 31.00 ± 10.61 | p=0.051 |
| ***Perceived stress (PSS)*** | 18.61 ± 7.66 | 12.25 ± 5.33 | p<0.01** |

### 3.2 Dynamic functional connectivity alterations in PPPD

#### rTPJ-seeded Co-activation patterns

When using the rTPJ as a seed region, 3 stable CAPs were identified. CAP1 showed co-activation of the rTPJ with salience/ventral attention network regions, and co-deactivation primarily with regions of the visual network but also with the dorsal attention network (**Fig.1a**). This CAP showed a significant decrease in patients with PPPD for both relative entries (pFDR<0.001) and relative occurrence (pFDR=0.009, **Fig.1b**), compared to HC. CAP2 is characterized by a co-activation of the rTPJ with regions of the visual, somatomotor, and salience networks, and co-deactivation with regions of the default mode and executive control networks. This CAP did not differ in its relative entries or relative occurrence between groups (all p>0.05). CAP3 shows a co-activation of the rTPJ with regions of the visual, dorsal attention, ventral attention and executive control networks, with co-deactivation with regions of the default mode network. This CAP showed a significant increase in relative entries in patients with PPPD compared to HC (pFDR=0.01), but not in relative occurrence (CAP3 pFDR=0.57). The average duration of rTPJ-seeded CAPs did not show any group differences (all p>0.05, **Suppl. Fig.2a**). Overall, rTPJ-seeded CAPs show an increase in total time or frequency of patterns coupled with regions of the visual and dorsal attention networks, and a decrease in patterns coupled with regions of the salience and default mode networks, in patients with PPPD compared to HC.

**Figure 1.**
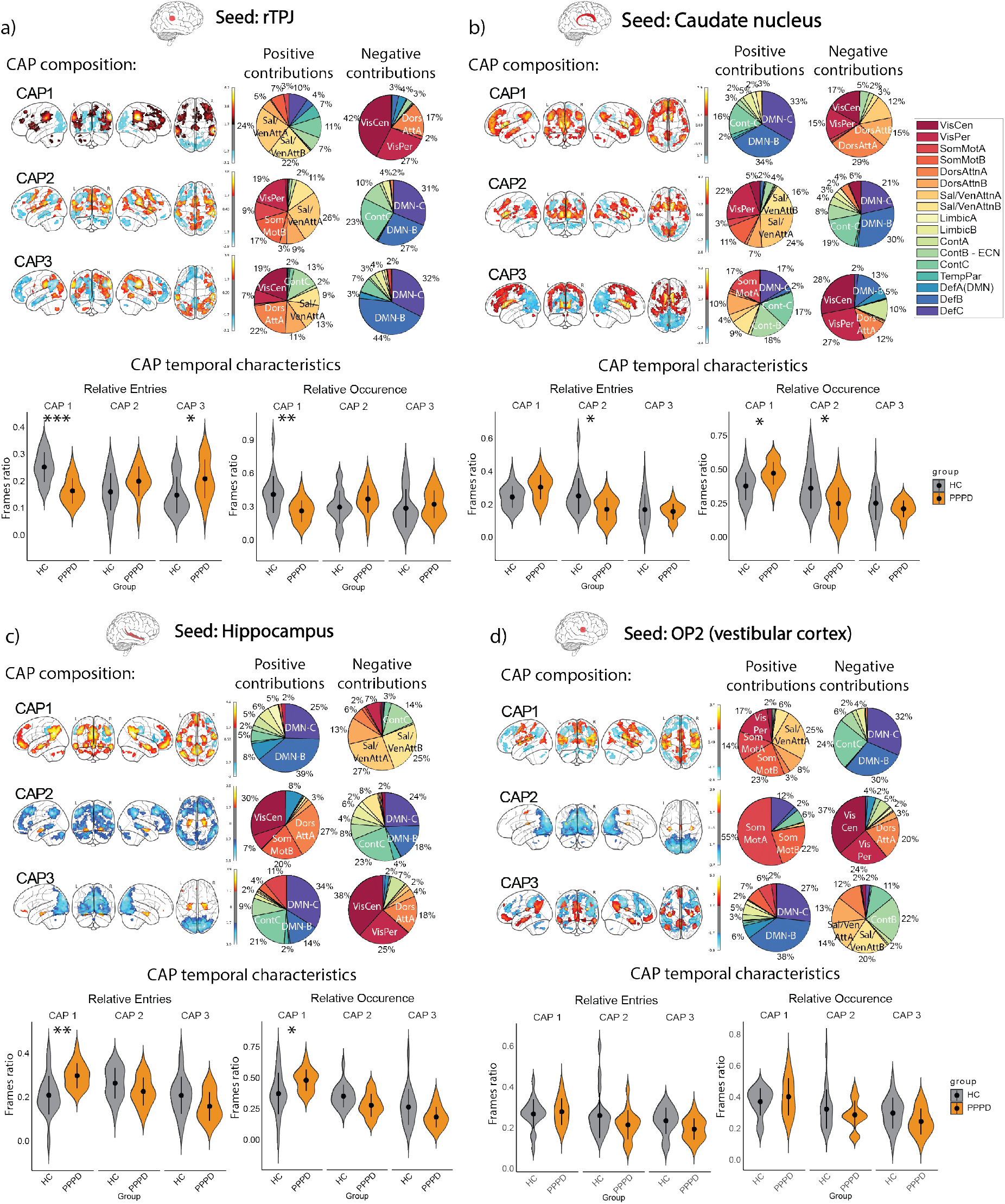
Co-Activation Patterns composition and temporal characteristics. CAPs were derived from HCs as reference population. In the CAP brain representations, red indicates co-activation (positive contributions, top 5%) and blue indicates co-deactivation (negative contributions, top 5%). The networks to which these regions map onto are shown in the pie charts, while the temporal characteristics of the CAPs are shown in the violin plots. **a)** Top: Co-Activation Patterns seeded from the rTPJ (right temporo-parietal junction). 3 CAPs were identified as stable clusters, with the first CAP showing co-activation of the rTPJ especially with regions of the salience/ventral attention network and co-deactivation with regions of the visual and dorsal attention networks. The second CAP shows co-activation of the rTPJ with regions of the visual, somatomotor, and salience networks, and co-deactivation with regions of the default mode and executive control networks. The third CAP shows co-activation of the rTPJ with regions of the visual, dorsal attention, ventral attention and executive control networks, with co-deactivation with regions of the default mode network. Bottom: temporal characteristics of rTPJ-seeded CAPs. In PPPD patients, CAP1 shows a significant decrease in relative entries and relative occurrence, while CAP3 shows a significant increase relative entries (relative entries: post-hoc comparison for CAP1 pFDR<0.001, Cohen’s d=-0.75; CAP2 pFDR=0.10, Cohen’s d=1.01, CAP3 pFDR=0.01, Cohen’s d=0.15; relative occurrence: post-hoc comparison for CAP1 pFDR=0.009, Cohen’s d=1.04; CAP2 pFDR=0.18, Cohen’s d=-0.51, CAP3 pFDR=0.57, Cohen’s d=-0.25; relative duration all p>0.05). **b)** Top: Co-Activation Patterns seeded from the caudate nucleus (bilateral). 3 CAPs were identified as stable clusters, with the first CAP showing co-activation of the caudate nucleus with regions of default mode and executive control networks and co-deactivation with regions of the visual and dorsal attention networks. The second CAP shows co-activation of the caudate nucleus with regions of the visual peripheral (i.e., early visual) and salience/ventral attention networks, and co-deactivation with regions of the default mode and executive control networks. The third CAP shows co-activation of the caudate nucleus with regions of the executive control, somatomotor and default mode A networks, with co-deactivation with regions of the visual, dorsal attention and default mode B network. Bottom: temporal characteristics of CAPs seeded from the caudate nucleus. In PPPD patients, CAP2 shows a significant decrease in relative entries and relative occurrence, and CAP1 shows a significant increase in relative occurrence. (relative entries: post-hoc comparison for CAP1 pFDR=0.054, Cohen’s d=-0.75; CAP2 pFDR=0.01, Cohen’s d=1.01, CAP3 pFDR=0.82, Cohen’s d=0.15; relative occurrence: post-hoc comparison for CAP1 pFDR=0.03, Cohen’s d=-0.85; CAP2 pFDR=0.01, Cohen’s d=1.0, CAP3 pFDR=0.40, Cohen’s d=0.36; relative duration all p>0.05). **c)** Top: Co-Activation Patterns seeded from the hippocampus (bilateral). 3 CAPs were identified as stable clusters, with the first CAP showing co-activation of the hippocampus with regions of the default mode network and co-deactivation with regions of the salience/ventral attention and executive control networks. The second CAP shows a prominent co-deactivation of the hippocampus with regions of the default mode and executive control networks. The third CAP shows a prominent co-deactivation of the hippocampus with regions of the visual network. Bottom: temporal characteristics of CAPs seeded from the hippocampus. In PPPD patients, CAP1 shows a significant increase in both relative entries and relative occurrence (relative entries: post-hoc comparison for CAP1 pFDR=0.003, Cohen’s d=-1.22; CAP2 pFDR=0.17, Cohen’s d=0.52, CAP3 pFDR=0.09, Cohen’s d=0.67; relative occurrence: post-hoc comparison for CAP1 pFDR=0.03, Cohen’s d=-0.91; CAP2 pFDR=0.11, Cohen’s d=0.63, CAP3 pFDR=0.09, Cohen’s d=0.69; relative duration all p>0.05). **d)** Top: Co-Activation Patterns seeded from the OP2 (opercular cortex 2, bilateral). 3 CAPs were identified as stable clusters, with the first CAP showing co-activation of the OP2 with regions of the visual, somatomotor, and salience networks and co-deactivation with regions of default mode and executive control networks. The second CAP shows a prominent co-deactivation with regions of the visual and dorsal attention networks. The third CAP shows co-activation of the OP2 with regions of the default mode network, with co-deactivation with regions of the salience and executive control networks. Bottom: temporal characteristics of CAPs seeded from the OP2. No significant difference was found in any CAP and characteristic between PPPD and HC groups (relative entries: all p>0.05; relative occurrence: all p>0.05; relative duration all p>0.05). For all results, * = 0.05>p≥0.01, ** = 0.01>p≥0.001, *** = p<0.001.

#### Caudate nucleus-seeded Co-activation patterns

When investigating the dynamic functional connectivity of the caudate nucleus, 3 stable CAPs were identified, with CAP1 showing co-activation with regions of the default mode and executive control networks and co-deactivation with regions of the visual and dorsal attention networks (**Fig.1c**). This CAP showed an increase in relative occurrence in patients with PPPD compared to HC (CAP2 pFDR=0.01), but not in relative entries (pFDR=0.054, **Fig.1d**). CAP2 shows co-activation of the caudate nucleus with regions of the visual peripheral (i.e., early visual cortex[44]) and salience/ventral attention networks, and co-deactivation with regions of the default mode and executive control networks. This pattern showed a decrease in both relative entries (pFDR=0.01) and relative occurrence (CAP2 pFDR=0.01) in patients with PPPD compared to HC. CAP3 shows co-activation of the caudate nucleus with regions of the executive control, somatomotor and default mode A networks, with co-deactivation with regions of the visual, dorsal attention and default mode B network. This CAP did not show significant differences in any temporal metric between groups. The average duration of caudate nucleus-seeded CAPs did not show any group differences (all p>0.05, **Suppl. Fig.2b**). Overall, caudate-seeded CAPs showed a decrease in patterns coupled with regions of the salience and dorsal attention networks, and an increase in patterns coupled with regions of the default mode network in patients with PPPD compared to HC. with regions of the default mode network.

#### Hippocampus-seeded Co-activation patterns

Three stable CAPs were also identified when using the hippocampus as a seed region. CAP1 showed co-activation of the hippocampus with regions of the default mode network and co-deactivation with regions of the salience/ventral attention and executive control networks (**Fig.1e**). This brain state was significantly higher for patients with PPPD, compared to HC, in both relative entries (pFDR=0.003) and relative occurrence (pFDR=0.03, **Fig.1f**). CAP2 showed prominent co-deactivation of the hippocampus with regions of the default mode and executive control networks, while CAP3 showed mostly a co-deactivation of the hippocampus with regions of the visual network. None of these two patterns showed group differences (all p>0.05). The average duration of all hippocampus-seeded CAPs did not show any group differences (all p>0.05, **Suppl. Fig.2c**). Overall, hippocampus-seeded CAPs show an increase in patterns coupled with the default mode network in patients with PPPD compared to HC.

#### OP2-seeded Co-activation patterns

We also obtained 3 stable CAPs seeded from the OP2 (operculo-parietal 2) cortex. CAP1 showed co-activation of the OP2 with regions of the visual, somatomotor, and salience networks and co-deactivation with regions of default mode and executive control networks. CAP2 showed a prominent co-deactivation with regions of the visual and dorsal attention networks. CAP3 showed a co-activation of the OP2 with regions of the default mode network and a co-deactivation with regions of the salience and executive control networks (**Fig.1g**). None of the CAPs seeded from the OP2 showed significant differences in their temporal characteristics between patients with PPPD and HC (all p>0.05, **Fig.1h** and **Suppl. Fig.2d**).

### 3.3 Relationship between dynamic functional connectivity and behavior

The multivariate PLSC analysis revealed two significant modes of brain-behavior coupling, or Latent Components (LCs), with distinct behavioral and imaging variables contributing to each mode. Note that, in the context of PLSC analysis, variables related to behavior but also to clinical assessment and biological factors are referred to as ‘behavioral variables’ for simplicity.

The first component, LC1 (p=0.013), explains 42% of covariance and is generated by behavioral variables related to biological and self-reported stress (salivary α-amylase and PSS), mood, vertigo, and gait velocity Δ (i.e. difference when walking with open and closed eyes, **Fig.2a**). Of note, the stress-related variables of patients with PPPD have a salience of the opposite sign of the other variables in this component. Additionally, the gait velocity Δ of the two groups has a salience of the same sign, indicating a similar contribution. Overall, the behavioral variables of LC1 are related to stress and gait characteristics. The imaging variables that generate LC1 are rTPJ-CAP1 (characterized by a co-activation with the salience network, and a positive contribution to LC1), OP2-CAP2 (characterized by a co-deactivation with the visual network and a negative contribution to the component), and Hippocampus-CAP3 (characterized by a co-activation with the default mode network and a negative contribution to LC1, **Fig.2b**). The LC1 component shows a positive correlation of behavioral and imaging scores at the individual level for both HC and PPPD groups (HC Spearman’s rho=0.56, p=0.022; PPPD Pearson’s r=0.74, p=0.001), indicating that the identified CAPs are related to behavior (**Suppl. Fig.3a**). The loadings of the behavioral and imaging variables resemble the pattern of saliences (**Suppl. Fig.3b-c**). Overall, this component relates stress and gait to brain CAPs originating from different seeds.

**Figure 2.**
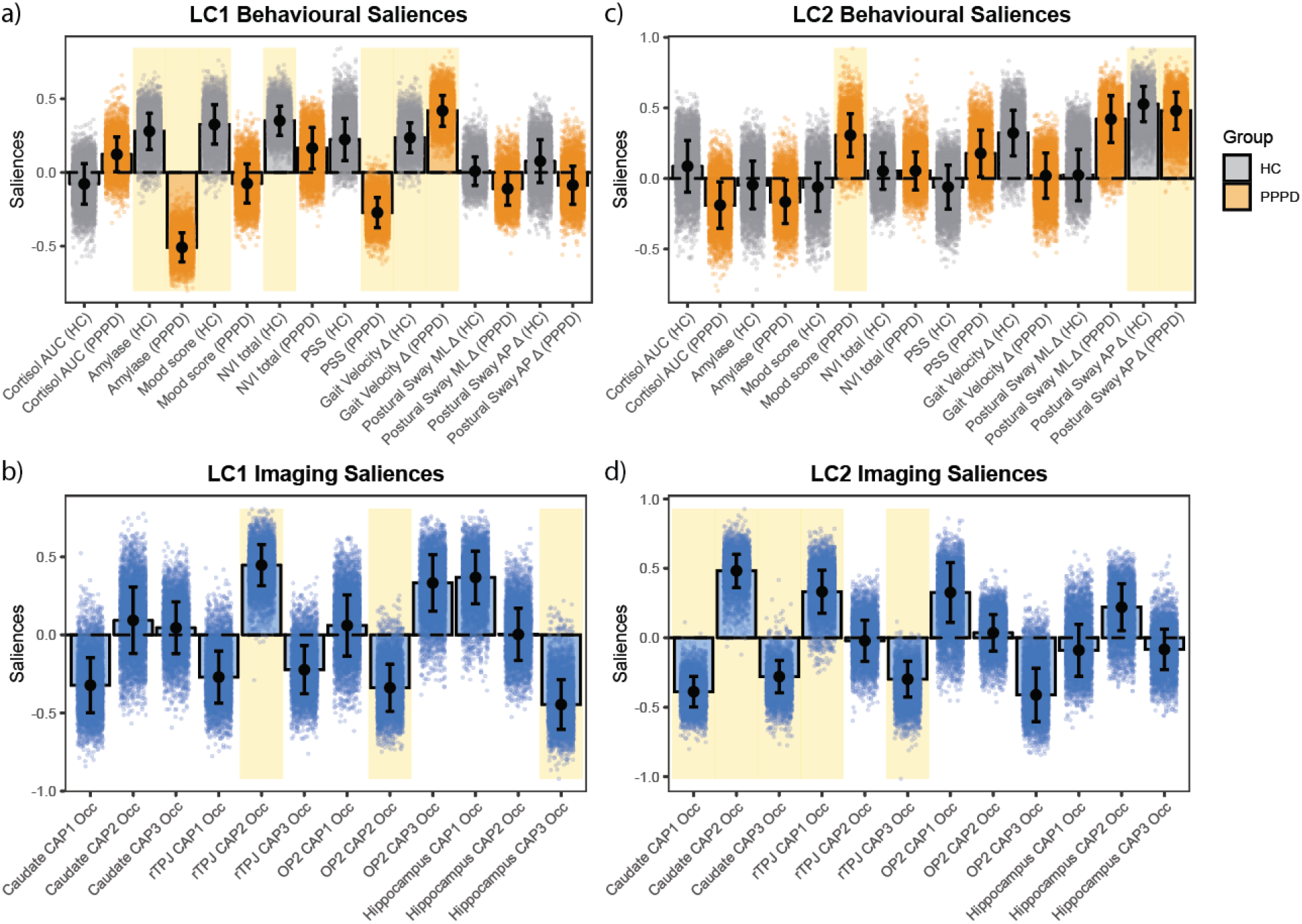
Significant latent components and their saliences for behavioral and imaging variables. Two significant brain-behavior modes of covariance (or Latent Components, LC) were identified through PLSC analysis, made up by different brain-behavior variables. Saliences indicate the variable’s contribution to the multivariate mode of covariance, i.e. how much each variable contributes to generating the mode. The error bars represent the standard deviation of the bootstrap distribution, and yellow shades represent variables whose salience weights were both statistically significant and robust, i.e. stable across bootstrap resamples (shaded dots). **a-b)** The first mode, LC1 (explained covariance = 42%, p=0.013), is generated by behavioral variables (a) related to mood, stress, vertigo, and gait metrics, and by the occurrence of CAPs (b) seeded from the rTPJ (CAP2), OP2 (CAP2), and Hippocampus (CAP3). **c-d)** The second mode, LC2 (explained covariance = 28%, p=0.016), is generated by behavioural variables (c) related to mood and postural sway, and by the occurrence (d) of the 3 caudate-seeded CAPs (CAP1, CAP2, CAP3) and 2 CAPs seeded from the rTPJ (CAP1 and CAP3).

The second significant component identified, LC2 (p=0.016), explains 28% of covariance, and is generated by the behavioral variable of mood for patients with PPPD, and postural sway Δ for both groups (**Fig.3c**). Of note, the salience of the postural variables of the same sign for both groups indicates a similar contribution. The imaging variables that constitute this component (**Fig.3d**) are all 3 CAPs seeded from the caudate nucleus, and CAP1 and CAP3 of the rTPJ (characterized by a co-activation with the salience network, and a co-deactivation with the default mode network, respectively). Like LC1, also this component shows a significant positive correlation of behavioral and imaging scores at the individual level for both HC and PPPD groups (HC Pearson’s r=0.64, p=0.006; PPPD Spearman’s rho=0.55, p=0.03), indicating that the identified CAPs are related to behavior (**Suppl. Fig.3d**) and the loadings resemble the pattern of saliences (**Suppl. Fig.3e-f**). Overall, this component relates postural sway to brain CAPs dominated by the caudate nucleus.

## 4. Discussion

We have employed a dynamic functional connectivity approach to identify alterations in the temporal expression of brain states in PPPD compared to HC at rest, and a multivariate correlation analysis to investigate their relationships with behavioral and clinical variables. Confirming our hypothesis, we have identified altered brain states in regions involved in multisensory integration (rTPJ), spatial navigation (hippocampus), and motor planning (caudate nucleus), but not in primary vestibular processing (OP2). We further uncovered that the dynamic alterations mostly related to postural control in this population are those seeded from motor and multisensory regions.

### 4.1 Brain dynamics seeded from multisensory and motor hubs, but not vestibular cortex, show altered temporal characteristics in patients with PPPD

#### rTPJ-seeded Co-activation patterns

Two rTPJ CAPs showed significantly different temporal dynamics between patients with PPPD and HC, with an overall decrease in a brain state coupled with the salience network (CAP1) and an increase in relative entries into a state coupled with the visual and dorsal attention networks (CAP3) in patients with PPPD. This might reflect an altered integration of salient stimuli, a greater reliance on vision and/or an increased top-down attentional control. Alterations in the TPJ have been found in PPPD by a recent study employing magnetoencephalography (MEG) and their low-frequency component was positively associated with dizziness symptoms[48]. Additionally, altered functional connectivity and hypoactivity of the rTPJ[26,49,50], as well as alterations in CAPs seeded from the rTPJ, have been previously reported in FND[42] Thus, our findings of altered rTPJ-CAPs underline the similarity of PPPD with the broader category of FND. The increase in rTPJ-seeded brain states coupled with the visual and dorsal attention networks might indicate a greater involvement of these regions in the multisensory integration of visual and vestibular cues, which contributes to the sense of self/agency. This echoes the fact that patients with PPPD are known to rely excessively on vision[11] and explicit attention[8,51] for postural control. This might be reflected in an increased coupling of the visual and dorsal attentional (explicit, goal-directed) networks with the rTPJ, accompanied by a decreased coupling with the salience network (stimulus-dependent, automatic attention), as seen here.

#### Caudate nucleus-seeded Co-activation patterns

Two CAPs seeded from the caudate nucleus showed significantly different temporal dynamics between groups, with increased relative occurrence, i.e., total time spent, in a state characterized by a positive coupling with the default mode network (CAP1) and decreased relative occurrence and entries of a brain state characterized by coupling of the caudate nucleus with the visual and salience networks (CAP2). The caudate nucleus, part of the basal ganglia, is a subcortical structure responsible for motor and cognitive functions such as the integration of spatial information with motor planning, and goal-directed action[27,28]. An increased coupling with the default mode network - associated with self-referential processing - might reflect an increased drawing from explicit self-related information for motor planning and execution. The decrease of a caudate-seeded brain state coupled with the salience network might instead reflect a reduced capacity to integrate salient external stimuli into motor commands, i.e. to adapt motor plans based on unexpected sensory changes, possibly underlying a disrupted coordination of postural signals. These findings underline how alterations can be found in patients with PPPD not just in cortical associative regions, but also in a subcortical structure responsible for coordinating motor output. This complement from the functional perspective a previous structural study which identified grey matter volume reductions in various brain regions in patients with PPPD, including the caudate nucleus[30].

#### Hippocampus-seeded Co-activation patterns

One CAP seeded from the hippocampus, characterized by coupling with the default mode network and de-coupling with the salience network, was found to be increased in temporal metrics of relative occurrence and entries in patients with PPPD. This finding might reflect an increased reliance on self-referential information for coordinating self-motion in space, a possible compensatory adaptation to counter the perceived loss of reliability in vestibular inputs and their multisensory integration with other sensory signals. Altered connectivity of the hippocampus in PPPD[31] has been hypothesized to underlie impaired spatial orientation and self-motion perception, a view supported by behavioral impairments in allocentric spatial navigation found in PPPD[52], a function which depends on the hippocampus. Additionally, an increased activation of the default mode network, has also been found in FND, linked to excessive self-monitoring[53], and its functional connectivity has been reported to be altered in PPPD[54].

#### OP2-seeded Co-activation patterns

In our sample, no differences have been found in CAPs seeded from the OP2 area. Thus, at rest, we do not find evidence of changes in the dynamic functional connectivity of the OP2 in patients with PPPD. This contrasts with the dynamic connectivity of other brain regions investigated here at the resting-state, and is compatible with previous literature on the disorder, which underscores that PPPD is not a primary vestibular dysfunction, but rather results from altered multisensory integration at the network level[7,10,12]. In line with our results, a previous study reported no differences in overall seed-based connectivity centered in the OP2 at rest, albeit with a decrease in functional connectivity between the left hippocampus and the left OP2[31], a feature that might not be picked up by CAPs analysis, which examines dynamic brain-wide states. These findings at rest raise the question of whether vestibular stimulation, or a task requiring active vestibular processing, might be required to uncover group differences in brain states seeded from this region. A recent study tested this by employing galvanic vestibular stimulation during fMRI scanning, revealing increased self-reported ego-motion upon stimulation in patients with PPPD compared to HC, accompanied by increased activation of multisensory vestibular areas such as OP3 and OP4, rather than OP2[34]..

Taken together, our findings support the absence of primary vestibular alterations in PPPD, and the presence of alterations in multisensory integration and motor output orchestration.

### 4.2 Two modes of brain-behavior organization map differently onto stress, mood, gait, and postural characteristics

Our PLSC analysis identified two distinct modes, or multi-covariance relationships, between the identified brain states (CAPs) and behavioral and clinical variables. Both these modes, or Latent Components, showed a significant positive correlation of individual brain and behavior scores – in other words, the higher an individual participant scored in the combination of CAPs that make up a component, the higher they scored also in the combination of behavioral variables belonging to that component. This is tested within each group after z-scoring of variables, thus removing the influence of pre-existing mean group differences, suggesting these modes represent meaningful brain-behavior relationships.

Interestingly, at the behavioral level, the two components showed a separation between stress and gait-related behavioral variables, which constitute LC1, and postural variables, which instead constitute LC2. Additionally, the two components also separated different brain variables (CAPs). The fact that both components showed significant positive correlations between individual brain and behavior scores in both groups underlines that these multivariate relationships identify a common physiological organization.

In LC1, we note the opposite contribution (i.e. salience of opposite sign) for salivary α-amylase of the two groups. While keeping in mind that the functional role of one variable cannot be isolated from the rest of the multivariate pattern, this suggests that this stress marker might contribute differently in the two groups. Additionally, the gait Δ variable, while showing a significant salience for both groups, presents a significant loading only in the patient group, i.e. an alignment with the latent pattern that is not observed in HC. Overall, this component is constituted by brain variables that did not reveal mean group differences in previous univariate analyses and by behavioral variables, as in the case of the combined mood score and the visual dependence in gait velocity (gait Δ), that are considered to be secondary to the pathophysiology of PPPD, i.e. a consequence of postural instability (also see[13]). This suggests that this component identifies a common pattern of brain-behavior organization, with slight differences on how this organization is present in the two groups.

The LC2 component, on the other hand, relates the postural sway Δ in the antero-posterior (AP) direction – an index of visual dependence for postural control in this axis – predominantly with CAPs seeded from the caudate nucleus and the rTPJ. The segregation of gait and postural variables into two distinct components is in line with the fact that patients with PPPD do not present primary gait alterations, but a cautious gait deriving from their primary postural disturbance. Since the caudate nucleus is a crucial region for motor planning, the fact that the multivariate pattern of LC2 links its altered dynamic connectivity with visual reliance for postural control suggests this might be a meaningful brain signature of this behavioral feature. Indeed, explicit attention is thought to exacerbate symptoms in both PPPD and FND[55]. Of note, the salience of the postural sway Δ has the same sign for both groups, and the same happens with its loading, suggesting that this brain-behavior relationship is present in both groups. Interestingly, CAPs seeded from the caudate nucleus and the rTPJ make up this component, suggesting that a synergy of these two regions might be important for postural control under visual challenge. Contrary to LC1, the brain and behavioral variables that constitute LC2 had previously shown mean group differences in univariate analyses, which is compatible with this result as the two analyses address different aspects of the data (mean differences and covariance within groups).

Together, this suggests that, although these brain and behavioral components are part of a common organizational relationship, they are differently expressed in patients with PPPD and HC. In other words, patients with PPPD do not seem to have a completely different brain-behavior organization, but rather an alteration in an existing organization, suggesting this might be re-trained with appropriate interventions.

## 5. Limitations

Despite efforts to follow current methodological best practices, this study presents limitations that need to be kept in mind when interpreting its findings. The sample size of this study was limited, therefore our findings might not generalize to the whole PPPD population. It was, however, in line with previous studies on this disorder[7]. Of note, our sample also differs slightly from previously reported sex ratios for PPPD, generally with a higher prevalence in females[55], and some participants in our sample reported intake of psychotropic medication, for which a subgroup analysis was not possible due to the small sample size. CAPs analysis was based on the *a priori* definition of seed regions based on previous literature and hypotheses, thus it should be kept in mind that it does not represent brain-wide, but rather seed-dependent, dynamic states. This note of caution transfers as well to our multivariate PLSC analysis, which thus represents brain-behavior relationships associated with the selected seeds. The results of CAPs analysis may depend on choices about specific analysis parameters (thresholding, clustering, and assignment threshold, use of PCA) and the use of the global signal regression for noise reduction in the preprocessing of fMRI data[56]. Additionally, although CAPs analyses has been successfully applied to identify transient patterns and their relationships to behavioral and task states in healthy populations[57], as well as to symptoms and their normalization after treatment in clinical populations[20,58], the exact neurobiological basis of these dynamic states is not yet fully elucidated, thus limiting inferences about function. Finally, the cross-sectional design of this study does not allow to test whether the found brain-behavior relationships are stable over time or change with symptom fluctuations. Finally, the cross-sectional design of this study does not allow to test whether the found brain-behavior relationships are stable over time or change with symptom fluctuations.

## 6. Conclusion

In this study, we have set out to characterize the brain dynamic functional connectivity patterns at rest in patients with PPPD and matched HC, and related these to clinically relevant self-reported and behavioral variables collected in the same sample. To our knowledge, this is the first study employing a dynamic connectivity approach in this population. We identify brain signatures of altered connectivity with the rest of the brain, seeded from brain regions previously reported to have an altered connectivity in the disorder, such as the hippocampus and the caudate nucleus, while also identifying alterations in other regions previously less investigated in the disorder, such as the rTPJ. Our results find no alterations, at rest, in the dynamic connectivity of the area OP2, identified as the primary vestibular cortex. These findings add and extend previous reports of functional and network alterations in PPPD predominantly in multimodal regions and regions involved in general motor and spatial planning over a disturbance of primary vestibular processing. Next, we identify two modes of organization (LCs) of brain and behavioral variables, distinctly related to postural control on one side, and to other variables on the other. Specifically, a component related postural control and affective symptoms with dynamic patterns derived from the caudate nucleus and the rTPJ. The fact that these modes of organization shared similar features across both groups suggests that the physiological brain-behavior organization is preserved but differently expressed in PPPD. Thus, we could speculate that a normalization of the dynamic alterations of PPPD might be achieved through focused vestibular rehabilitation, which targets visual dependence, or cognitive-behavioral therapy with attenion-focused approaches. If confirmed in larger samples, some of the dynamic brain signatures identified here might serve in the future as endpoints to track the reduction of symptoms and of visual dependence for postural control in PPPD.

## 7. Author contributions

**CRediT author contributions**: Cristina Concetti: *Conceptualization, Methodology, Data Curation, Formal analysis, Project administration, Visualization, Writing – original draft, Writing – review and editing,*; Anna Wyss: *Conceptualization, Methodology, Project administration, Investigation, Data curation;* Irene Lozzi: *Data Curation*; Salome Häuselmann: *Methodology, Data Curation, Project administration, Investigation*; Tatiana Brémovà-Ertl: *Resources*; Hassen Kerkeni: *Methodology, Resources*; Georgios Mantokoudis: *Resources*, Nicolas Gninenko: *Supervision, Writing – review and editing*, Michael Mouthon: *Writing – review and editing*, Maria Giulia Preti: *Supervision;* Dimitri Van De Ville: *Supervision*; Selma Aybek: *Conceptualization, Resources, Supervision, Funding acquisition*.

## 8. Acknowledgements

We thank the patients and their close ones as well as the healthy volunteers who made this study possible with their participation.

## Data Availability

The full dataset used for the current study cannot be shared due to ethical and privacy restrictions.

