## Supplementary Materials for "Brain Dynamics of Persistent Postural-Perceptual Dizziness at Rest Reveal Alterations in Multisensory and Motor Areas"

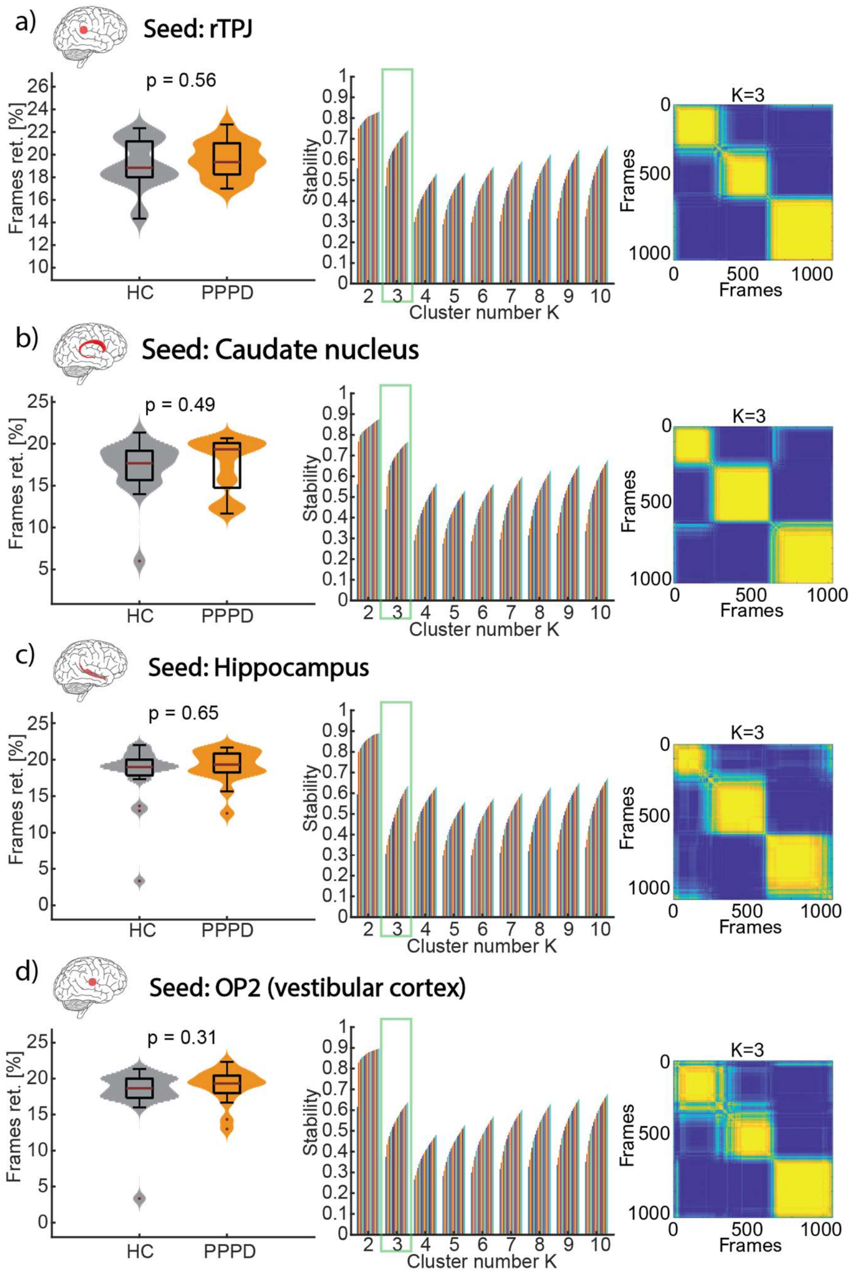


**Supplementary Figure 1. CAPs analysis Quality Control metrics (retained frames, cluster stability and consensus matrix).** For each seed region, the left panel shows the number of frames retained for analysis based on the selection of highly active frames (above 0.84 SD of the z-scored time series) of the seed ROI. For each seed, the number of frames retained for clustering did not differ significantly across groups: a) rTPJ p=0.56, b) Caudate nucleus p=0.49, c) Hippocampus p=0.65, d) OP2 p=0.31, Student's t-test). The middle panel shows cluster stability across different cluster numbers (K, or CAPs) using the k-means algorithm. Stability is defined as 1-PAC (Percentage of Ambiguously Clustered pairs). For each seed, the cluster number >2 with the highest stability was identified as the optimal number of CAPs. The right panel shows the consensus clustering matrix for the K number (number of CAPs) identified as stable, where the yellow color and sharp edges between clusters indicate for each frame that it is repeatedly assigned to the same cluster.

| | **HC (mean ± SD)** | **PPPD (mean ± SD)** |
|---|---|---|
| ***rTPJ*** | | |
| Retained frames | 57.26 ± 6.46 | 48.76 ± 4.76 |
| Excluded (scrubbed) frames | 2.25 ± 4.61 | 2.70 ± 5.77 |
| Unassigned frames | n.a. | 3.74 ± 3.64 |
| ***Caudate*** | | |
| Retained frames | 51,55 ± 9.52 | 52.35 ± 9.61 |
| Excluded (scrubbed) frames | 1.8 ± 4.3 | 2 ± 3.27 |
| Unassigned frames | n.a. | 5.82 ± 4.24 |
| ***Hippocampus*** | | |
| Retained frames | 54.15 ± 12 | 57.23 ± 6.78 |
| Excluded (scrubbed) frames | 2.15 ± 4.35 | 2.23 ± 3.74 |
| Unassigned frames | n.a. | 4.73 ± 3.75 |
| ***OP2*** | | |
| Retained frames | 54.05 ± 10.97 | 56.82 ± 7.01 |
| Excluded (scrubbed) frames | 1.75 ± 3.88 | 2.47 ± 4.64 |
| Unassigned frames | n.a. | 5.89 ± 4.24 |

**Supplementary Table 1. Additional CAPs analysis Quality Control metrics.** For each seed, the absolute number of frames retained for CAPs analysis is reported, as well as the absolute number of excluded frames (scrubbed due to motion>0.5 mm) and the absolute number of unassigned frames for the PPPD group (assignment index Ap=5%).

| | Positive contributions | | | Negative contributions | | |
|---|---|---|---|---|---|---|
| | Peak MNI coordinates (x, y, z) | Corresponding regions | | Peak MNI coordinates (x, y, z) | Corresponding regions | |
| | | AAL Atlas | Schaefer 200 Atlas | | AAL Atlas | Schaefer 200 Atlas |
| ***rTPJ*** | | | | | | |
| CAP 1 | 62, -36, 32 | SupraMarginal_R | RH_Cont_Par_1 | 8, -86, -12 | Lingual_R | RH_Vis_6 |
| | -60, -36, 30 | SupraMarginal_L | LH_SalVentAttn_ParOper_3 | -24, -76, -14 | Fusiform_L | LH_Vis_2 |
| | 38, 48, 22 | Frontal_Mid_2_R | RH_Cont_PFCl_3 | 24, -40, -14 | Fusiform_R | RH_Vis_2 |
| | 8, -30, 46 | Cingulate_Mid_R | RH_SalVentAttn_Med_2 | 6, -60, 12 | Calcarine_R | RH_Default_pCunPCC_1 |
| | -2, 32, 52 | Frontal_Sup_Medial_L | LH_Default_PFC_10 | 10, -46, 2 | Lingual_R | RH_Default_pCunPCC_1 |
| | -54, 8, 6 | Frontal_Inf_Oper_L | LH_SalVentAttn_FrOperIns_4 | 46, -70, -16 | Occipital_Inf_R | RH_DorsAttn_Post_1 |
| | -38, 42, 22 | Frontal_Mid_2_L | LH_Cont_PFCl_3 | 6, -88, 8 | Calcarine_R | RH_Vis_9 |
| | -20, -80, -48 | Cerebelum_Crus2_L | Background | -30, -90, 14 | Occipital_Mid_L | LH_Vis_11 |
| | 58, -24, -6 | Temporal_Mid_R | RH_Default_Temp_4 | 26, -78, 40 | Occipital_Sup_R | RH_Vis_15 |
| | 4, 34, 50 | Frontal_Sup_Medial_R | RH_Cont_PFCmp_2 | | | |
| | -6, 12, 36 | Cingulate_Mid_L | LH_SalVentAttn_Med_1 | | | |
| | -40, -54, -30 | Cerebelum_Crus1_L | Background | | | |
| | 12, 4, 70 | Supp_Motor_Area_R | RH_SalVentAttn_Med_3 | | | |
| | -38, -52, -44 | Cerebelum_7b_L | Background | | | |
| | 4, 12, 42 | Cingulate_Mid_R | RH_SalVentAttn_Med_1 | | | |
| | -32, 26, 38 | Frontal_Mid_2_L | LH_SalVentAttn_PFCl_1 | | | |
| CAP 2 | 58, -32, 32 | SupraMarginal_R | RH_SalVentAttn_TempOccPar_3 | 22, 38, 52 | Frontal_Sup_2_R | RH_Default_PFCdPFCm_5 |
| | 4, 12, 42 | Cingulate_Mid_R | RH_SalVentAttn_Med_1 | 38, 16, 54 | Frontal_Mid_2_R | RH_Cont_PFCl_7 |
| | -60, -32, 22 | Temporal_Sup_L | LH_SalVentAttn_ParOper_2 | -10, 62, 30 | Frontal_Sup_Medial_L | LH_Default_PFC_7 |
| | -8, -70, 10 | Calcarine_L | LH_Vis_10 | -10, -84, -26 | Cerebelum_Crus2_L | Background |
| | 12, -82, 40 | Cuneus_R | RH_Vis_14 | -12, 68, 14 | Frontal_Sup_2_L | LH_Default_PFC_7 |
| | 12, -30, 44 | Cingulate_Mid_R | RH_SalVentAttn_Med_2 | 64, -4, -22 | Temporal_Mid_R | RH_Default_Temp_2 |
| | -6, -48, 62 | Precuneus_L | LH_DorsAttn_Post_9 | -2, -52, 22 | Cingulate_Post_L | LH_Default_pCunPCC_2 |
| | 32, 46, 26 | Frontal_Mid_2_R | RH_Cont_PFCl_5 | 64, -38, -8 | Temporal_Mid_R | RH_Cont_Temp_1 |
| | 52, -4, 48 | Precentral_R | RH_SomMot_10 | -44, -72, -38 | Cerebelum_Crus2_L | Background |
| | -22, -48, 72 | Parietal_Sup_L | LH_DorsAttn_Post_10 | 12, -84, -26 | Cerebelum_Crus1_R | Background |
| | -32, 36, 28 | Frontal_Mid_2_L | LH_SalVentAttn_PFCl_1 | 46, 24, 44 | Frontal_Mid_2_R | RH_Cont_PFCl_7 |
| | 48, 40, 4 | Frontal_Inf_Tri_R | RH_Cont_PFCl_3 | -44, 52, -2 | Frontal_Mid_2_L | LH_Cont_PFCl_1 |
| | -18, -66, -16 | Cerebelum_6_L | Background | -2, 54, -18 | Rectus_L | LH_Default_PFC_4 |
| | | | | 42, -68, -42 | Cerebelum_Crus2_R | Background |
| | | | | 46, -66, 44 | Angular_R | RH_Default_Par_3 |
| | | | | -36, 22, 54 | Frontal_Mid_2_L | LH_Default_PFC_11 |

| | | | | | | |
|---|---|---|---|---|---|---|
| | | | | -64, -34, -6 | Temporal_Mid_L | LH_Cont_Temp_1 |
| | | | | -38, -66, 54 | Parietal_Sup_L | LH_Cont_Par_2 |
| CAP 3 | 60, -30, 32 | SupraMarginal_R | RH_SalVentAttn_TempOccPar_3 | 44, -72, -36 | Cerebelum_Crus1_R | Background |
| | -60, -34, 32 | SupraMarginal_L | LH_SalVentAttn_ParOper_3 | 24, 38, 44 | Frontal_Sup_2_R | RH_Default_PFCdPFCm_6 |
| | 56, -54, -4 | Temporal_Inf_R | RH_DorsAttn_Post_1 | 60, -2, -24 | Temporal_Mid_R | RH_Default_Temp_2 |
| | -52, -64, -2 | Temporal_Mid_L | LH_DorsAttn_Post_2 | -62, -10, -18 | Temporal_Mid_L | LH_Default_Temp_2 |
| | 52, 12, 26 | Frontal_Inf_Oper_R | RH_DorsAttn_PrCv_1 | -2, -52, 40 | Precuneus_L | LH_Default_pCunPCC_4 |
| | -42, -82, 16 | Occipital_Mid_L | LH_Vis_11 | -48, -70, 36 | Angular_L | LH_Default_Par_4 |
| | -24, -74, -50 | Cerebelum_7b_L | Background | -2, 54, -10 | Frontal_Med_Orb_L | LH_Default_PFC_4 |
| | 46, 40, 6 | Frontal_Inf_Tri_R | RH_Cont_PFCl_3 | 52, -62, 32 | Angular_R | RH_Default_Par_1 |
| | -18, -66, 58 | Parietal_Sup_L | LH_DorsAttn_Post_7 | | | |
| | -46, 4, 28 | Frontal_Inf_Oper_L | LH_DorsAttn_PrCv_1 | | | |
| | 38, 50, 28 | Frontal_Mid_2_R | RH_Cont_PFCl_5 | | | |
| | -54, 8, 12 | Frontal_Inf_Oper_L | LH_SalVentAttn_FrOperIns_4 | | | |
| | 10, -34, 46 | Cingulate_Mid_R | RH_SalVentAttn_Med_2 | | | |
| | -14, -76, 44 | Parietal_Sup_L | LH_Cont_pCun_1 | | | |
| | -32, -52, 56 | Parietal_Sup_L | LH_DorsAttn_Post_6 | | | |
| ***Caudate*** | | | | | | |
| CAP 1 | 10, 14, 8 | Caudate_R | Background | 32, -80, 26 | Occipital_Mid_R | RH_Vis_12 |
| | -8, 16, 2 | Caudate_L | Background | -20, -64, 56 | Parietal_Sup_L | LH_DorsAttn_Post_7 |
| | -2, 56, -10 | Frontal_Med_Orb_L | LH_Default_PFC_4 | -10, -80, 48 | Precuneus_L | LH_Cont_pCun_1 |
| | -48, -66, 40 | Angular_L | LH_Default_Par_4 | 44, -74, 16 | Occipital_Mid_R | RH_Vis_11 |
| | 52, -62, 36 | Angular_R | RH_Default_Par_3 | -62, -30, 32 | SupraMarginal_L | LH_SalVentAttn_ParOper_2 |
| | -62, -26, -8 | Temporal_Mid_L | LH_Default_Temp_4 | -2, -82, 40 | Cuneus_L | LH_Vis_13 |
| | -4, -54, 38 | Precuneus_L | LH_Default_pCunPCC_4 | 58, -30, 34 | SupraMarginal_R | RH_SalVentAttn_TempOccPar_3 |
| | 60, -4, -28 | Temporal_Mid_R | RH_Default_Temp_2 | 14, -84, 42 | Cuneus_R | RH_Vis_14 |
| | 40, 32, -14 | OFCpost_R | RH_Cont_PFCl_1 | | | |
| | 42, -74, -38 | Cerebelum_Crus2_R | Background | | | |
| | -54, 24, 18 | Frontal_Inf_Tri_L | LH_Default_PFC_5 | | | |
| | -48, 28, -10 | Frontal_Inf_Orb_2_L | LH_Default_PFC_3 | | | |
| | -4, -58, -48 | Cerebelum_9_L | Background | | | |
| | 42, 24, 44 | Frontal_Mid_2_R | RH_Cont_PFCl_7 | | | |
| | -32, 18, -18 | Insula_L | LH_Default_PFC_1 | | | |
| | -22, -86, -36 | Cerebelum_Crus2_L | Background | | | |
| | 24, -80, -28 | Cerebelum_Crus1_R | Background | | | |
| | -36, 52, -2 | Frontal_Mid_2_L | LH_Cont_PFCl_1 | | | |
| CAP 2 | 10, 6, 6 | Caudate_R | Background | 2, 62, -6 | Frontal_Med_Orb_R | RH_Limbic_OFC_3 |
| | -8, 6, 4 | Caudate_L | Background | 62, -40, -8 | Temporal_Mid_R | RH_Cont_Temp_1 |

| | | | | | | |
|---|---|---|---|---|---|---|
| | 4, 16, 44 | Cingulate_Mid_R | RH_SalVentAttn_Med_1 | 64, -12, -20 | Temporal_Mid_R | RH_Default_Temp_2 |
| | 12, -76, 12 | Calcarine_R | RH_Vis_9 | -50, -64, -20 | no_label | LH_DorsAttn_Post_1 |
| | 58, -34, 36 | SupraMarginal_R | RH_SalVentAttn_TempOccPar_3 | -44, 12, 54 | Frontal_Mid_2_L | LH_Default_PFC_11 |
| | 44, 0, 50 | Precentral_R | RH_DorsAttn_FEF_1 | -60, -40, -10 | Temporal_Mid_L | LH_Cont_Temp_1 |
| | -36, 48, 20 | Frontal_Mid_2_L | LH_Cont_PFCl_3 | 48, -70, -28 | Cerebelum_Crus1_R | Background |
| | -58, -18, 26 | Postcentral_L | LH_SalVentAttn_ParOper_2 | 2, -56, 24 | Precuneus_R | RH_Default_pCunPCC_2 |
| | 50, 36, 4 | Frontal_Inf_Tri_R | RH_Cont_PFCl_3 | -30, -76, 48 | Parietal_Inf_L | LH_Cont_Par_2 |
| | 34, 46, 22 | Frontal_Mid_2_R | RH_Cont_PFCl_5 | -20, -92, -30 | Cerebelum_Crus2_L | Background |
| | -42, -4, 52 | Precentral_L | LH_DorsAttn_FEF_1 | 38, -68, 48 | Angular_R | RH_Cont_Par_3 |
| | -36, -16, -2 | Insula_L | LH_SalVentAttn_FrOperIns_1 | | | |
| | 12, -26, 42 | Cingulate_Mid_R | RH_SalVentAttn_Med_2 | | | |
| | -28, 34, -14 | OFCpost_L | LH_Limbic_OFC_1 | | | |
| | -14, -80, -8 | Lingual_L | LH_Vis_2 | | | |
| | -56, -16, 10 | Temporal_Sup_L | LH_SomMot_2 | | | |
| | -18, -56, -6 | Lingual_L | LH_Vis_1 | | | |
| CAP 3 | -8, 12, 12 | Caudate_L | Background | 36, -56, -18 | Fusiform_R | RH_Vis_3 |
| | -56, 18, 24 | Frontal_Inf_Tri_L | LH_Default_PFC_5 | 28, -80, -8 | Fusiform_R | RH_Vis_3 |
| | 56, 18, 30 | Frontal_Inf_Oper_R | RH_Cont_PFCl_4 | 48, -60, 12 | Temporal_Mid_R | RH_DorsAttn_Post_2 |
| | 6, -4, 14 | no_label | Background | -24, -48, -10 | Lingual_L | LH_Vis_1 |
| | -42, -42, 56 | Parietal_Sup_L | LH_DorsAttn_Post_5 | 50, -70, 0 | Temporal_Mid_R | RH_Vis_5 |
| | -4, -8, 16 | no_label | Background | 4, -72, 36 | Precuneus_R | RH_Cont_pCun_1 |
| | 42, -40, 58 | Postcentral_R | RH_DorsAttn_Post_4 | | | |
| | -6, -82, -26 | Cerebelum_Crus2_L | Background | | | |
| | 6, -84, -26 | Cerebelum_Crus2_R | Background | | | |
| | 4, 38, 54 | Frontal_Sup_Medial_R | RH_Cont_PFCmp_2 | | | |
| | -32, 60, 14 | Frontal_Mid_2_L | LH_Cont_PFCl_2 | | | |
| | 0, -58, -2 | Vermis_4_5 | Background | | | |
| | -18, 46, 44 | Frontal_Sup_2_L | LH_Default_PFC_9 | | | |
| ***Hippocampus*** | | | | | | |
| CAP 1 | 4, -52, 22 | Precuneus_R | RH_Default_pCunPCC_2 | 50, 42, 2 | Frontal_Inf_Tri_R | RH_Cont_PFCl_3 |
| | 60, -6, -20 | Temporal_Mid_R | RH_Default_Temp_2 | 34, -42, 40 | no_label | RH_DorsAttn_Post_4 |
| | 2, 52, -14 | Frontal_Med_Orb_R | RH_Default_PFCdPFCm_1 | 10, -32, 48 | Paracentral_Lobule_R | RH_SalVentAttn_Med_2 |
| | 26, -20, -16 | Hippocampus_R | Background | 46, 16, -4 | Insula_R | RH_SalVentAttn_FrOperIns_4 |
| | 52, -62, 32 | Angular_R | RH_Default_Par_1 | 38, 50, 24 | Frontal_Mid_2_R | RH_Cont_PFCl_5 |
| | -42, -72, 40 | Angular_L | LH_Default_Par_4 | -58, -36, 32 | SupraMarginal_L | LH_SalVentAttn_ParOper_3 |
| | -60, -12, -16 | Temporal_Mid_L | LH_Default_Temp_3 | 62, -30, 32 | SupraMarginal_R | RH_SalVentAttn_TempOccPar_3 |
| | -42, 12, -36 | Temporal_Pole_Mid_L | LH_Default_Temp_1 | | | |

| | | | | | | |
|---|---|---|---|---|---|---|
| | 22, 40, 46 | Frontal_Sup_2_R | RH_Default_PFCdPFCm_5 | | | |
| | -20, 38, 50 | Frontal_Sup_2_L | LH_Default_PFC_12 | | | |
| | -12, -88, -40 | Cerebelum_Crus2_L | Background | | | |
| | 44, -68, -38 | Cerebelum_Crus1_R | Background | | | |
| | -6, -54, -44 | Cerebelum_9_L | Background | | | |
| | 16, -90, -34 | Cerebelum_Crus2_R | Background | | | |
| | 40, 34, -14 | OFCant_R | RH_Cont_PFCl_1 | | | |
| CAP 2 | 24, -16, -16 | Hippocampus_R | Background | 34, 20, -14 | Insula_R | RH_Limbic_OFC_2 |
| | -22, -18, -16 | Hippocampus_L | Background | 2, -22, 42 | Cingulate_Mid_R | RH_Cont_Cing_1 |
| | | | | 16, -102, -6 | Calcarine_R | RH_Vis_8 |
| | | | | -10, 56, -24 | OFCmed_L | LH_Limbic_OFC_2 |
| | | | | 0, -70, 44 | Precuneus_L | LH_Default_pCunPCC_4 |
| | | | | 42, -76, -42 | Cerebelum_Crus2_R | Background |
| | | | | 52, 24, 8 | Frontal_Inf_Tri_R | RH_Default_PFCv_1 |
| | | | | -42, 20, -16 | Temporal_Pole_Sup_L | LH_Default_PFC_1 |
| | | | | -56, 22, 8 | Frontal_Inf_Tri_L | LH_Default_PFC_5 |
| | | | | -66, -52, -2 | Temporal_Mid_L | LH_DorsAttn_Post_2 |
| | | | | -38, 56, -10 | Frontal_Mid_2_L | LH_Cont_PFCl_1 |
| | | | | -16, -84, -30 | Cerebelum_Crus1_L | Background |
| | | | | 68, -38, -6 | Temporal_Mid_R | RH_Cont_Temp_1 |
| | | | | -58, -52, 40 | Parietal_Inf_L | LH_Cont_Par_1 |
| | | | | -44, 16, 48 | Frontal_Mid_2_L | LH_Default_PFC_11 |
| | | | | 46, 22, 42 | Frontal_Mid_2_R | RH_Cont_PFCl_7 |
| | | | | 58, -48, 40 | Parietal_Inf_R | RH_Default_Par_2 |
| CAP 3 | 22, -10, -16 | Hippocampus_R | Background | 0, -60, -38 | Vermis_9 | Background |
| | -22, -22, -16 | Hippocampus_L | Background | 4, -82, 36 | no_label | RH_Vis_14 |
| | -50, 38, -10 | Frontal_Inf_Orb_2_L | LH_Default_PFC_3 | | | |
| | -2, 44, -8 | Frontal_Med_Orb_L | LH_Default_PFC_2 | | | |
| ***OP2*** | | | | | | |
| CAP 1 | 34, -24, 18 | Insula_R | RH_SomMot_3 | 64, -32, -12 | Temporal_Mid_R | RH_Cont_Temp_1 |
| | -34, -26, 18 | Rolandic_Oper_L | LH_SomMot_3 | 62, -6, -26 | Temporal_Mid_R | RH_Default_Temp_2 |
| | 6, 10, 44 | Cingulate_Mid_R | RH_SalVentAttn_Med_1 | -10, -84, -28 | Cerebelum_Crus2_L | Background |
| | -10, -42, 56 | Precuneus_L | LH_SomMot_15 | -42, -72, -42 | Cerebelum_Crus2_L | Background |
| | 18, -64, 8 | Calcarine_R | RH_Vis_10 | 8, -84, -26 | Cerebelum_Crus2_R | Background |
| | 14, -82, 40 | Cuneus_R | RH_Vis_14 | 22, 66, 10 | Frontal_Sup_2_R | RH_Cont_PFCl_2 |
| | -38, -18, 40 | Postcentral_L | LH_SomMot_7 | -42, 56, -4 | Frontal_Mid_2_L | LH_Cont_PFCl_1 |

| | | | | | | |
|---|---|---|---|---|---|---|
| | 52, -4, 48 | Precentral_R | RH_SomMot_10 | -20, 66, 10 | Frontal_Sup_2_L | LH_Default_PFC_4 |
| | -38, -20, 0 | Insula_L | LH_SalVentAttn_FrOperIns_1 | 0, -62, 40 | Precuneus_L | LH_Default_pCunPCC_4 |
| | -26, -46, 70 | Parietal_Sup_L | LH_SomMot_11 | -2, 54, -18 | Rectus_L | LH_Default_PFC_4 |
| | | | | 40, -78, -44 | Cerebelum_Crus2_R | Background |
| | | | | -64, -30, -10 | Temporal_Mid_L | LH_Cont_Temp_1 |
| | | | | 48, 22, 42 | Frontal_Mid_2_R | RH_Cont_PFCl_7 |
| | | | | -22, 34, 52 | Frontal_Sup_2_L | LH_Default_PFC_12 |
| | | | | 4, 54, 40 | Frontal_Sup_Medial_R | RH_Default_PFCdPFCm_4 |
| | | | | 52, -60, 34 | Angular_R | RH_Default_Par_3 |
| | | | | -38, -64, 54 | Parietal_Inf_L | LH_Cont_Par_2 |
| CAP 2 | 34, -22, 16 | Insula_R | RH_SomMot_3 | -18, -62, 4 | Calcarine_L | LH_Vis_10 |
| | -34, -24, 18 | Insula_L | LH_SomMot_3 | 18, -42, -46 | Cerebelum_9_R | Background |
| | 2, -18, 54 | Supp_Motor_Area_R | RH_SomMot_18 | 0, -60, -40 | Vermis_9 | Background |
| | | | | -20, -42, -46 | Cerebelum_9_L | Background |
| | | | | -8, -80, 48 | Precuneus_L | LH_Cont_pCun_1 |
| CAP 3 | 36, -24, 18 | Rolandic_Oper_R | RH_SomMot_4 | 22, -78, -50 | Cerebelum_7b_R | Background |
| | -36, -24, 18 | Rolandic_Oper_L | LH_SomMot_3 | -48, 6, 26 | Frontal_Inf_Oper_L | LH_DorsAttn_PrCv_1 |
| | 62, -6, -22 | Temporal_Mid_R | RH_Default_Temp_2 | -32, -2, 46 | Precentral_L | LH_DorsAttn_FEF_1 |
| | 2, -54, 26 | Precuneus_R | Background | -2, 20, 48 | Supp_Motor_Area_L | LH_Default_PFC_10 |
| | -62, -8, -20 | Temporal_Mid_L | LH_Default_Temp_2 | 10, -30, 44 | Cingulate_Mid_R | RH_SalVentAttn_Med_2 |
| | 2, 48, -16 | Rectus_R | RH_Default_PFCdPFCm_1 | -18, -66, 58 | Parietal_Sup_L | LH_DorsAttn_Post_7 |
| | -46, -64, 30 | Angular_L | LH_Default_Par_4 | -32, 22, 6 | Insula_L | LH_SalVentAttn_FrOperIns_2 |
| | 50, -60, 30 | Angular_R | RH_Default_Par_1 | -44, -58, -32 | Cerebelum_Crus1_L | Background |
| | -24, -22, -16 | Hippocampus_L | Background | 12, -72, 54 | Precuneus_R | RH_DorsAttn_Post_6 |
| | 26, -20, -16 | Hippocampus_R | Background | -58, -56, -4 | Temporal_Mid_L | LH_DorsAttn_Post_2 |
| | -12, 56, 32 | Frontal_Sup_2_L | LH_Default_PFC_9 | 4, 28, 42 | Frontal_Sup_Medial_R | RH_Cont_PFCmp_2 |
| | 42, 16, -32 | Temporal_Pole_Mid_R | RH_Default_Temp_1 | 62, -52, -6 | Temporal_Mid_R | RH_Cont_Temp_1 |
| | | | | -44, 48, 14 | Frontal_Mid_2_L | LH_Cont_PFCl_3 |
| | | | | 56, 16, 2 | Frontal_Inf_Oper_R | RH_Default_PFCv_1 |
| | | | | 62, -28, 42 | SupraMarginal_R | RH_SalVentAttn_TempOccPar_3 |
| | | | | -60, -36, 32 | SupraMarginal_L | LH_SalVentAttn_ParOper_3 |
| | | | | 40, 48, 26 | Frontal_Mid_2_R | RH_Cont_PFCl_5 |

**Supplementary Table 2. Brain regions contributing to CAPs**. For each seed and each CAP, the contributing regions are reported here as cluster peaks with their coordinates in the MNI space (x,y,z) and their mapping onto the anatomical AAL atlas as well as the functional Schaefer200 atlas, which is closely related to the Yeo functional network parcellation but offers a higher level of detail.

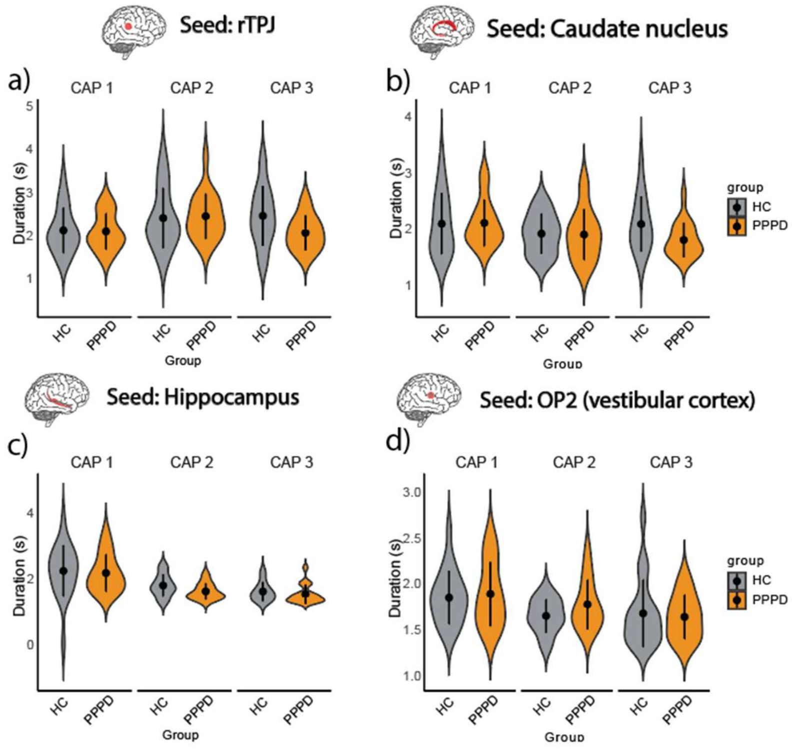


**Supplementary Figure 2. CAP duration.** a) Average duration of CAPs seeded from the rTPJ; b) from the caudate nucleus; c) from the hippocampus; d) from the OP2 area. The average duration of CAPs did not show any group differences (no main effect of group, no group x CAP interaction effect, all $p>0.05$).

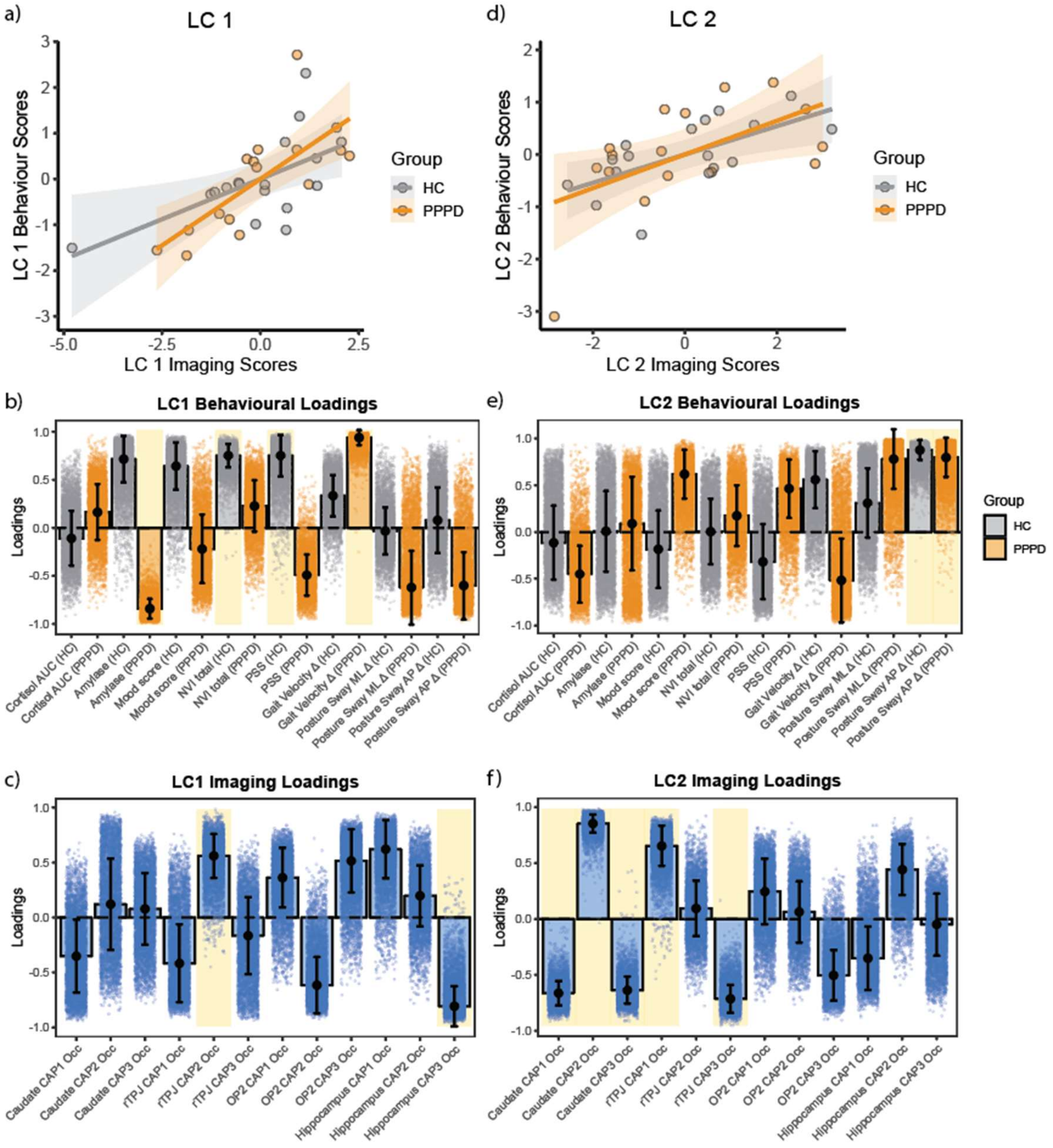


**Supplementary Figure 3. Brain-behavior correlations and loadings of significant latent components.** Panels a) and d) show brain-behavior correlations at the individual level for the identified significant latent components (LC). Panels b-f show the loadings of behavioral and imaging variables for each significant latent component. Loadings can be interpreted similarly to correlation scores or weights obtained when the LC is projected back into the underlying variables. The error bars represent the standard deviation of the bootstrap distribution, and yellow shades represent variables whose loadings were both statistically significant and robust, i.e. stable across bootstrap resamples (shaded dots). **a)** The first significant mode of brain-behaviour coupling, LC1, shows a positive correlation of behavioral and imaging scores at the individual level for both HC and PPPD groups (HC Spearman's rho=0.56, p=0.022; PPPD Pearson's r=0.74, p=0.001), indicating that the identified CAPs are related to behavioral variables. **b-c)** The behavioral and imaging loadings of LC1 closely resemble the saliences of behavioral and imaging variables of this component (Fig.2a-b), with significant behavioral variables (b) related to mood, stress, vertigo, and gait metrics, and the occurrence of CAPs (c) seeded from the rTPJ (CAP2), and Hippocampus (CAP3). **d)** The second significant mode of brain-behavior coupling, LC2, shows a positive correlation of behavioral and imaging scores at the individual level for both HC and PPPD groups (HC Pearson's r=0.64, p=0.006; PPPD Spearman's rho=0.55, p=0.03). **e-f)** The behavioral and imaging loadings of LC2 closely resemble the saliences of behavioral and imaging variables of this component (Fig.2c-d), with significant behavioural variables (e) related to postural sway, and the occurrence (f) of the 3 caudate-seeded CAPs (CAP1, CAP2, CAP3) and 2 CAPs seeded from the rTPJ (CAP1 and CAP3).
